# Personal and Contextual components of Resilience mediate Risky Family Environment’s effect on Psychotic-Like Experiences

**DOI:** 10.1101/2020.05.18.20106237

**Authors:** Rodolfo Rossi, Alberto Collazzoni, Dalila Talevi, Dino Gibertoni, Eleonora Quarta, Alessandro Rossi, Paolo Stratta, Giorgio Di Lorenzo, Francesca Pacitti

**Author notes:** **Correspondence:** Rodolfo Rossi, Department of Systems Medicine, University of Rome Tor Vergata, Via Montpellier 1, 00133, Roma, Italy.

## Abstract

**Background:** Psychotic-like experiences index an increased risk for subsequent psychotic disorders. A risky family environment is a well-established risk factor for psychotic-like experiences, however different contextual and personal factors may differentially mediate their effect on psychotic-like experiences, including different aspects of resilience.

**Objective:** In this study we propose a two-dimension model of resilience. Our aim is to address separately the mediational role of personal and contextual resilience factors between a risky family environment and PLE in a community sample.

**Methods and Materials:** Five hundred University students completed an on-line questionnaire including the Resilience Scale for Adults, the 16-item version of the Prodromal Questionnaire and the Risky Family Questionnaire.

Mediation was assessed using Structural Equation Modelling with bootstrapping estimation of indirect effect.

**Results:** Direct effect of Personal and Contextual resilience on Psychotic-like experiences were respectively −0.69 [−0.97, −0.41] (p<0.001) and −0.19 [−0.58, 0.20] (ns); indirect effects through personal resilience was 0.03[ 0.01, 0.04] (p<0.001). Personal resilience mediated 27.4% of the total effect of risky family environment on psychotic-like experiences.

**Discussion:** Personal resilience, but not contextual resilience, mediated the effect of a risky family environment on Psychotic-like experiences. Poor personal resilience may represent an individual risk factor that transmits the effect of risky family environment on psychotic-like experiences, and could represent a central aspect of individualized prevention and treatment strategies.

## Background

Psychotic-Like Experiences (PLEs) are sub-clinical psychotic phenomena relatively common in the general population, with an estimated lifetime prevalence of about 7% (Linscott & Van Os, 2013; McGrath et al., 2015), peaking during early adolescence and settling to a plateau in early adulthood (Thapar et al., 2012; Zammit et al., 2013). PLEs are conceptualized within the proneness-persistence-impairment model of psychotic disorders: individuals with a genetically-determined vulnerability to psychosis exposed to a number of risk factors, gradually increase the risk of having PLE and eventually transitioning to a psychotic disorder going through a prodromal at-risk mental state, although the exact rate of conversion is still a matter of debate. Such model is founded on the evidence that PLE and Psychotic Disorders share the same risk factors, including perinatal (Glaser et al., 2010), developmental (Thomas et al., 2009), neuropsychological (Horwood et al., 2008; R. Rossi et al., 2016) and emotional factors.

Childhood adversities, including a familial environment characterized by violence, unreliable or chaotic parenting style and neglect, are among the most well-established and replicated risk factors for all the stages of the psychosis continuum (Morgan & Gayer-Anderson, 2016). On the one side of the psychosis spectrum, childhood adversities are associated with PLE (Fisher et al., 2013; McGrath et al., 2017, 2015). On the other extreme, a recent meta-analysis has established a significant association between childhood adversities and psychosis (Varese et al., 2012) with odd ratios of 2.78. Between these two extremes, childhood adversity has been associated with persistence of psychotic experiences (Trotta, Murray, & Fisher, 2015) and Ultra-high risk (UHR) states (Peh, Rapisarda, & Lee, 2019).

The mechanisms through which childhood adversities and a risky family environment convey a heightened risk for PLE and psychotic disorders are still a matter of research. One of the leading lines of research is focusing on the putative *psychological* and *cognitive* mediators of the impact of childhood adversities on the psychotic continuum. To name a few, source monitoring deficits (Serrone et al., 2019) and dissociation (Bentall et al., 2014) are well-known mediators between sexual abuse and auditory hallucinations; reasoning biases mediate the development of paranoia (Freeman & Garety, 2014); and affective states (anxiety, depressive symptoms, external locus of control, and low self-esteem) have been shown to mediate the development of PLE (Fisher et al., 2013). An extensive systematic review by Williams and colleagues (Williams, Bucci, Berry, & Varese, 2018) has classified mediators in post-traumatic sequelae, affective disturbance and dysregulation, cognitive processes and appraisal of subsequent stressors.

Among the psychological mediators of potential interest for the psychotic continuum, resilience has received large attention.

Resilience is the capacity of adaptively overcoming stress and adversity while maintaining normal, or developing a better psychological functioning.

Resilience is a broad multimodal construct (Stainton et al., 2018) that comprises different personal and contextual aspects. Personal factors, also termed *assets*, include individual characteristics such as competence, coping skills, and self-efficacy. Contextual factors, or *resources*, are positive factors that are external to the individual, and include parental support, social connectedness or community organizations (Fergus & Zimmerman, 2005).

Resilience has been shown to be associated with risk factors for PLEs. For example, social support has shown a protective effect to PLEs against childhood adversity (Crush et al., 2018). On the other hand, some individual assets of resilience, such as stress sensitivity or self-confidence may be negatively affected by childhood adversity in the first place (Rauschenberg et al., 2017). When assessing subclinical outcomes in general population samples, resilience has been shown to mediate the effect of childhood adversities on PLE (Sengutta, Gawęda, Moritz, & Karow, 2019).

In clinical samples, resilience has been linked to different outcomes: for example, in psychotic patients low resilience has been related to depressive symptoms (A. Rossi et al., 2017) conveying suicidal risk to depressed patients (Rossetti et al., 2017), while higher levels of resilience participate in the process of personal recovery (A. Rossi et al., 2018) in patients with schizophrenia.

Under a psychometric perspective, a bi-factorial structure of the RSA has been recently confirmed using CFA in its Spanish version (Morote, Hjemdal, Uribe, & Corveleyn, 2017). Separating ‘contextual’ interpersonal from intrapersonal resilience resources may help elucidating some psychological mediational mechanisms underlying the effect of childhood adversity risky family environment on PLEs.

In the operationalization behind the Resilience Scale for Adults (RSA) (Friborg, Barlaug, Martinussen, Rosenvinge, & Hjemdal, 2005; Friborg, Hjemdal, Rosenvinge, & Martinussen, 2003), both personal and social resources are proposed. This distinction is somewhat similar to the definition of resilience-related interpersonal resources and individual assets proposed by Fergus and Zimmermann (Fergus & Zimmerman, 2005).

Following up the call to conceptualize resilience as a multimodal construct, the aim of this study is to examine an higher-order bi-factorial model of resilience made up of personal assets and interpersonal resources, and to separately address the mediational role of personal and interpersonal resilience factors between childhood adversity and PLE in a community sample. In particular, we test the hypothesis that exposure to a risky family environment could differentially affect resilience that in turns could exert a weakened protective effect on PLEs.

## Materials and Methods

### Participants

Participants were all bachelor and master’s degree students at University of L’Aquila. The recruitment was conducted online with the help of ads on different social networks related to the University. All participants provided written informed consent and the study was approved by the local ethics committee. Institutional Review Board code of this study is 22562/21.05.18

In order to ensure the validity participants’ answers, six “attention checks” were distributed throughout the entire survey, asking participants to answer in a particular way (i.e. “please answer “yes” to this question”). Two thousand one hundred and sixty-seven volunteers visited the online survey, but only five hundred gave the consent, answered correctly all of the attention checks and, filled in all of the questionnaires.

## Measures

### Risky Family Questionnaire

Childhood Adversity was assessed using the Risky Family Questionnaire (RFQ), a 13-items retrospective self-report questionnaire on a 5-point likert scale derived from the Adverse Childhood Experiences questionnaire (ACE-q) by Felitti et al., (Felitti et al., 1998). RFQ investigates the exposure to harsh parenting during childhood. Examples of the items are: “Would you say the household you grew up in was chaotic and disorganized?” and “Would you say you were neglected while you were growing up, left on your own to fend for yourself?”. Because the RFQ lacks of an Italian validation, we performed a split-sample Exploratory Factor Analysis (EFA) and a Confirmatory Factor Analysis (CFA) of the RFQ (supplementary materials).

### Resilience Scale for Adults

Resilience was assessed using the Resilience Scale for Adults (RSA), Italian version (RSA) (Capanna, Stratta, Hjemdal, Collazzoni, & Rossi, 2015; Friborg et al., 2003). The RSA is a 7-point Likert scale of 33 items grouped into 6 factors, namely *Perception of Self*: concerning self-confidence and positive outlook; *Planned Future:* concerning a positive outlook on one’s own future; *Social Competence:* concerning individual’s own perception of her/his ability to initiate verbal contact and flexibility in social interactions; *Structured Style:* concerning goal oriented planning ability; *Family Cohesion:* concerning shared values and cohesion within one’s family; *Social Resources:* concerning social support and feeling of cohesion outside the family. Psychometric details of the Italian version of the RSA can be found elsewhere (Capanna et al., 2015). In this sample, alpha coefficient was 0.92.

### Prodromal Questionnaire-16, Italian version

The Italian version of the Prodromal Questionnaire-16 (iPQ-16) (Azzali et al., 2018) was used to assess the presence of PLEs. iPQ-16 is a 16-itmes self-report instrument that explores the presence/absence of 16 PLEs, including perceptual aberrations/hallucinations, unusual thought content/delusions, and two negative symptoms, and their associated psychological distress. iPQ-16 scores the number of actual PLE endorsed, ranging from 0 to 16, and a distress score on a 4-point likert scale ranging from 0 to 48. Although the iPQ-16 was originally designed as a screening tool for individuals at UHR in help-seeking populations, several studies have used this instrument in the general population as a measure of PLEs (Gawęda, Göritz, & Moritz, 2019; Gawęda, Pionke, et al., 2019; Mętel et al., 2020; Sengutta et al., 2019). Because different cut-off points have been proposed for different populations (Savill, D’Ambrosio, Cannon, & Loewy, 2018), we chose to use the iPQ-16 score as a continuous rather than binary variable in order to avoid sensibility/specificity issues. For the sake of clarity of exposure, we will present only the results of the iPQ-16 endorsed score, as the results of the distress score were substantially overlapping.

## Statistical analysis

All the following statistics were performed using Stata 13^®^.

Firstly, descriptive statistics were performed on demographic variables, RSA, RFQ and iPQ-16.

Secondly, Confirmatory Factor Analysis (CFA) of RSA with Variance-Covariance matrix Maximum Likelihood estimation (ML) was performed on two theory-driven models: a one-dimensional solution, with the six factors loading on a single second order latent variable, and a two-dimension solution with *“Perception Of Self”*, “*Planned Future”, “Social Competence”*and *“Structured Style”* loading on one latent variable named *“PERSONAL RESILIENCE*’, and *“Family Cohesion*” and *“Social Resources*” loading on a second latent variable named *“CONTEXTUAL RESILIENCE*”. The two models have been contrasted (Morote et al., 2017) in a Spanish-language version, with better fit indices for the two-dimension model. To the best of our knowledge, no direct comparison of the two alternative models has been reported in a non-clinical Italian sample. Goodness-of-fit indices were computed for the two models, including relative fit indices Aikake’s Information Criteria (AIC) and Bayesian Information Criteria (BIC) that allow goodness of fit comparison between models.

Once a two-dimension structure of RSA was confirmed, a mediation analysis was performed within a SEM framework. RFQ total score and iPQ-16 total score were modelled, respectively, as exogenous and response variables. The two latent factors of resilience, Personal and Contextual Resilience, were included as simultaneous mediators. Observed scores of the six resilience factors, computed from the RSA questionnaire were used as manifest indicators of both latent variables for the sake of simplicity of calculation.

An initial model including all possible paths connecting exogenous variable, mediators and the response was specified. This model was further modified according to modification indices.

Mediation was assessed inspecting the indirect effect percentile and bias-corrected 95% confidence interval, according to Hayes and Preacher method. Hayes and Preacher bootstrapping method is currently considered the best method for assessing significance of the indirect effects (Shrout & Bolger, 2002).

## Results

### Characteristics of the sample

Demographic characteristics of the sample are shown in **table 1**. Five hundred participants completed the questionnaire, of which 356 (71.2%) were female. Mean age in the sample was 25.52 (SD=5.84). Summary statistics for RFQ, RSA and iPQ16 are reported in table 1.

**Table 1.** Demographic characteristics of the sample and descriptive statistics of variables of interest.

|  |  |
| --- | --- |
| **Age** |  |
| Range | 18-62 |
| Mean (SD) | 25.52 (5.84) |
| **Gender, n(%)** |  |
| Female | 356 (71.2%) |
| Male | 144 (28.80%) |
| **Education (years)** |  |
| Range | 8-30 |
| Mean (SD) | 17.35 (2.91) |
| **RFQ** |  |
| Range | 13-61 |
| Mean (SD) | 26.54 (8.68) |
| **RSA, mean (SD)** |  |
| *Perception Of The Self* | 4.69 (1.28) |
| *Planned Future* | 4.70 (1.51) |
| *Social Competence* | 4.88 (1.23) |
| *Family Cohesion* | 4.92 (1.53) |
| *Social Resources* | 5.63 (1.19) |
| *Structured Style* | 5.12 (1.22) |
| **iPQ-16 Endorsed score,** |  |
| mean (SD) | 4.02 (3.06) |

### RFQ Exploratory and Confirmatory Factor analysis

EFA performed on 250 randomly selected participants indicate a single-factor structure with 80% of variance explained, factor loadings comprised between 0.80 and 0.25 for the 10 positive items, and between −0.54 and −0.80 for the 3 reversed-scored items. Reliability □ coefficient was 0.87 with 0.39 average interitem covariance. CFA initially showed unsatisfactory fit indices. After inspection of modification indices, covariances between items were added accordingly. Fit indices in the resulting model were satisfactory, with RMSEA=0.083, CFI=0.91 and CD=0.87.

### CFA of RSA

CFA fit indices for the unidimensional and two-dimension proposed models are reported in **Table 2**. A graphical representation of the two-dimension model is presented in **Figure 1**. Both models showed adequate fit, and did not require post-hoc re-specification based on modification indices. Overall, the two-dimension model displayed slightly better fit indices, with lower AIC and BIC. Reaching convergence was particularly difficult for the two-dimension model and specification of starting values from a simplified model was necessary.

**Table 2.** Goodness-of-fit indices of Confirmatory Factor Analysis of RSA.

| Fit Index | One-dimensional | Two-dimension |
| --- | --- | --- |
| $\chi^2_{(489)}$ | 1310.217 | 1236.808 |
| p | 0.000 | 0.000 |
| RMSEA | 0.058 [0.054, 0.062] | 0.055 [0.052, 0.059] |
| AIC | 58920.746 | 58847.337 |
| BIC | 59363.070 | 59289.660 |
| CFI | 0.891 | 0.900 |
| SRMR | 0.088 | 0.139 |
| CD | 0.886 | 0.986 |

RSA: Resilience Scale for Adults, Italian version; RMSEA: Root mean squared error of approximation; AIC: Akaike's information criterion; BIC: Bayesian information criterion; CFI: Comparative fit index; SRMR: Standardized root mean squared residual; CD: Coefficient of determination.

**Figure 1:**
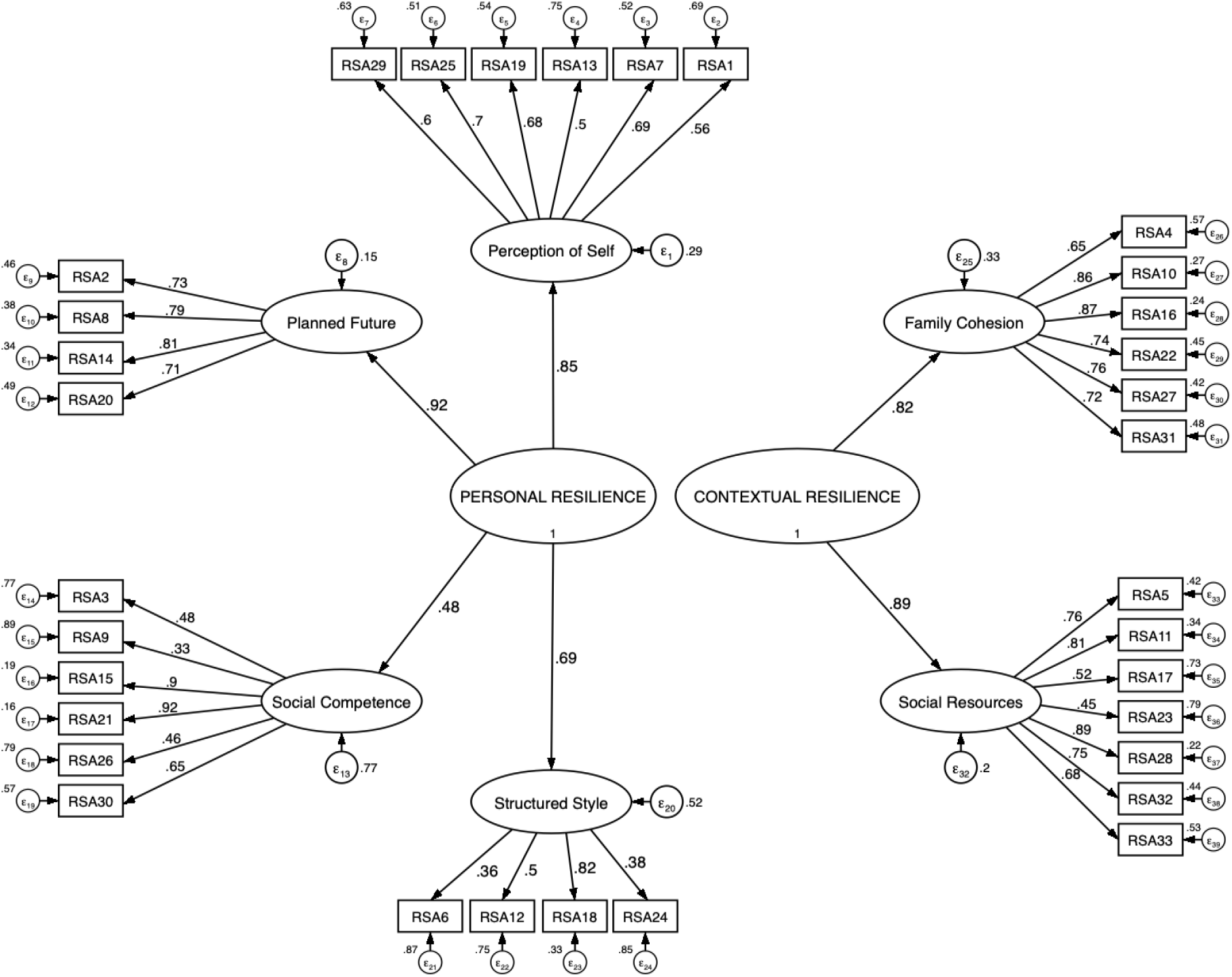
Confirmatory factor analysis of RSA with standardized coefficients.

### SEM and mediation

**Figure 2** reports the path diagram with standardized coefficients. All path coefficients were statistically significant except for the direct effect of Contextual Resilience on iPQ-16. After the initial model specification and inspection of the modification indices, a covariance between *Social Resources* and *Social Competence* was added. The resulting goodness of fit indices were as follows: 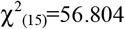 (p<0.001); RMSEA=0.075 [0.055, 0.096]; CFI=0.968; SRMR=0.040; CD=0.544. The estimates of direct, indirect and total effects are reported in **Table 3**. Because the direct effect between Contextual Resilience and iPQ-16 was not significant, we did not go further in assessing mediation through this path. The indirect effect between RFQ and iPQ-16 through Personal Resilience was 0.028 [0.013, 0.044]. As the total effect of RFQ on iPQ-16 was 0.1 [0.07, 0.13], 48.7% of the effect of RFQ on iPQ-16 was mediated by RSA as a whole, with 27.4% through Personal Resilience.

**Table 3.**
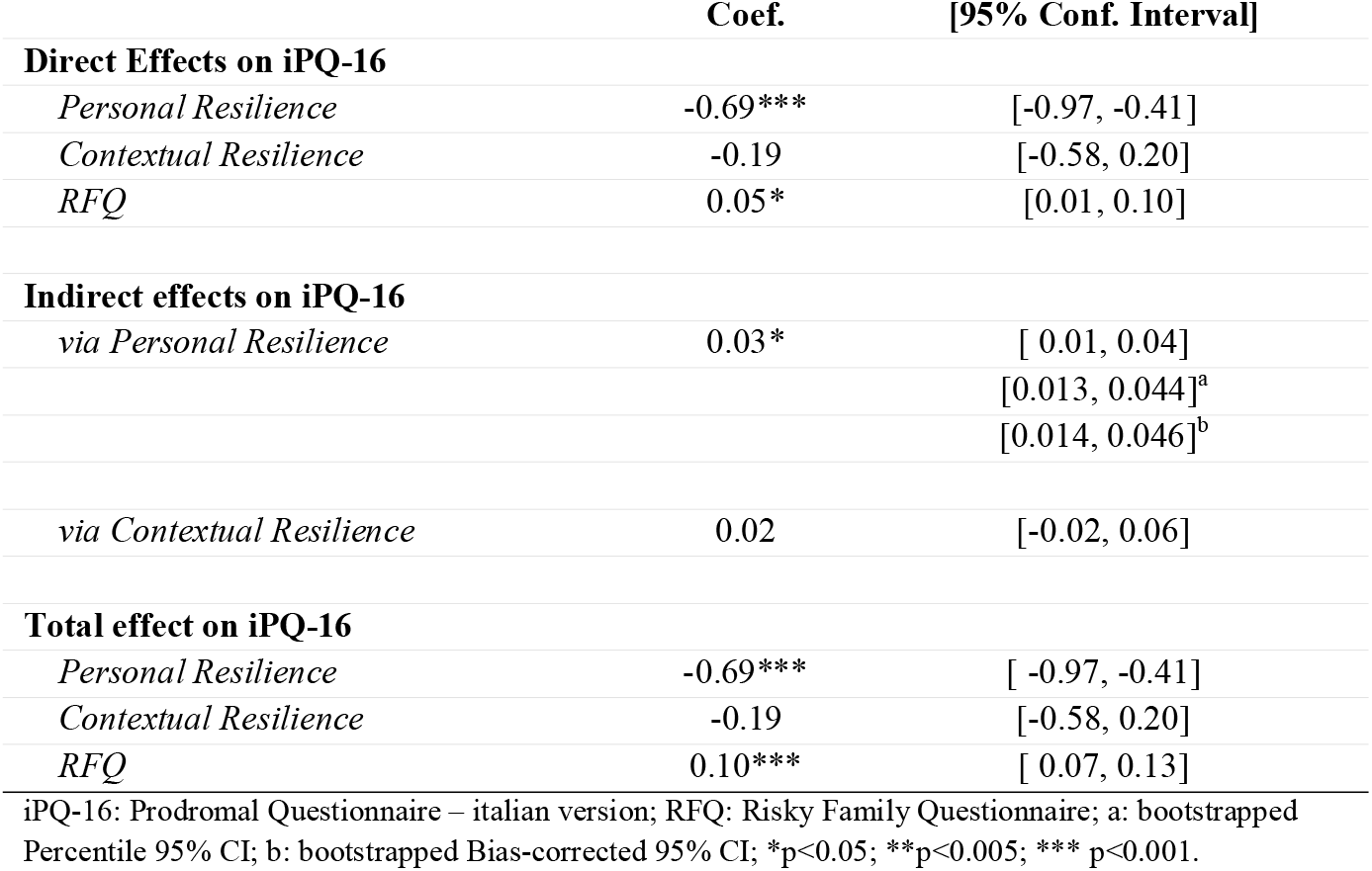
Path model summary with direct, indirect and total effects.

|  | Coef. | [95% Conf. Interval] |
| --- | --- | --- |
| **Direct Effects on iPQ-16** |  |  |
| *Personal Resilience* | -0.69*** | [-0.97, -0.41] |
| *Contextual Resilience* | -0.19 | [-0.58, 0.20] |
| *RFQ* | 0.05* | [0.01, 0.10] |
| **Indirect effects on iPQ-16** |  |  |
| *via Personal Resilience* | 0.03* | [ 0.01, 0.04] |
|  |  | [0.013, 0.044]<sup>a</sup> |
|  |  | [0.014, 0.046]<sup>b</sup> |
| *via Contextual Resilience* | 0.02 | [-0.02, 0.06] |
| **Total effect on iPQ-16** |  |  |
| *Personal Resilience* | -0.69*** | [ -0.97, -0.41] |
| *Contextual Resilience* | -0.19 | [-0.58, 0.20] |
| *RFQ* | 0.10*** | [ 0.07, 0.13] |

iPQ-16: Prodromal Questionnaire – italian version; RFQ: Risky Family Questionnaire; a: bootstrapped Percentile 95% CI; b: bootstrapped Bias-corrected 95% CI; \*p<0.05; \*\*p<0.005; \*\*\* p<0.001.

**Figure 2.**
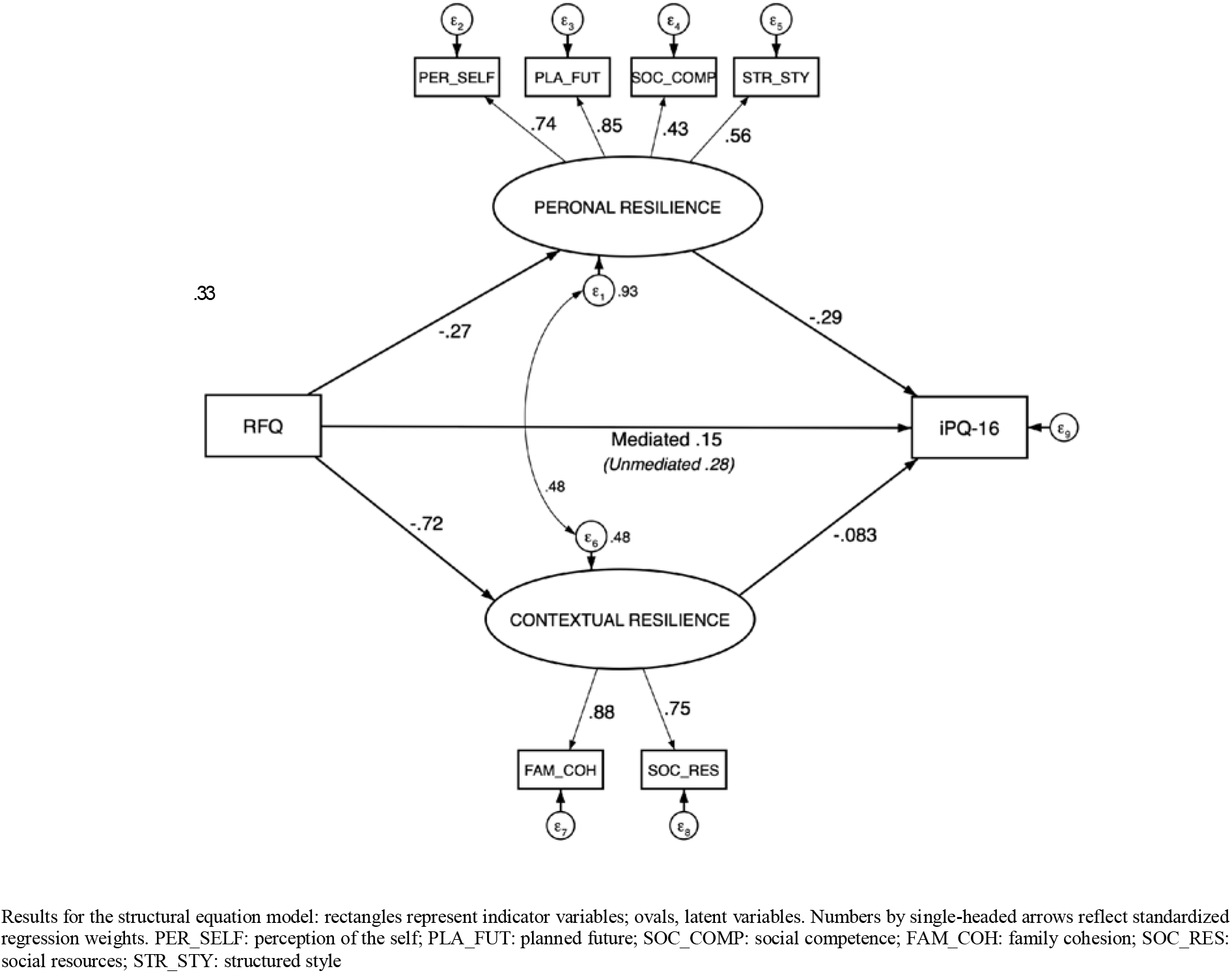
Path model with standardized coefficients.

We further tested the significance of the indirect effect using bias-corrected and percentile bootstrapped 95%CI inspection. Unstandardized indirect effects were computed for each of 1000 bootstrapped samples, and the 95% confidence interval was computed by determining the indirect effects at the 2.5th and 97.5th percentiles. The bootstrapped Percentile and Bias-corrected 95% CI of the indirect effect through Personal Resilience were respectively [0.013, 0.044] and [0.014, 0.046]. Thus, the indirect effect via Personal Resilience was statistically significant.

## Discussion

### Summary of findings

In the present study, we analyzed in detail the mediational effect of resilience between risky family environment and PLEs in a relatively large sample of university students.

In particular, we successfully decomposed resilience as measured by the RSA into two separated components using CFA: a personal component representing individual resiliency assets, and a contextual component representing resilience resources associated with social and familial aspects. Our results show that, although a unidimensional model adequately fits the data, a bidimensional model fits the data in a better way. Our finding is consistent with one previous CFA on a Spanish-speaking sample (Morote et al., 2017). Furthermore, a recent network analysis supports a four domain model with personal competence being the central node between “support” (family cohesion + social resources), “structured style” and “social competence”, a finding that is consistent with our CFA (Briganti & Linkowski, 2019). Overall, our findings are in line with the conceptualization of resilience proposed by Fergus and Zimmerman that clearly separate personal assets from social and interpersonal resources (Fergus & Zimmerman, 2005).

Next, we found evidence that personal and contextual domains of resilience have substantially different roles relative to risky family environment and PLEs, with nearly 28% of the effect of risky family on PLEs being mediated through personal resilience factors. Risky family environment had a substantial impact on contextual resilience, however no effect from contextual resilience to PLEs was observed in our model.

### Previous literature

Mediators between childhood adversity, including a risky family environment, and psychosis risk are a central topic in current research. Although resilience has been previously investigated as a key psychological factor for psychosis risk and psychosis outcome (Mętel et al., 2020; Ruzibiza, Grattan, Eder, & Linscott, 2018), this is the first study that addresses in detail the differential role of two separate components of resilience as risk factors for PLEs. The main narrative in current research conceptualizes resilience as a rather stable protective factor against childhood adversity and poor mental health outcomes (Southwick, Bonanno, Masten, Panter-Brick, & Yehuda, 2014). According to this view, resilience should be modelled as a potential moderator, rather than a mediator, of the effect of stressful or traumatic events on psychotic risk. This hypothesis wasn’t confirmed in a rather small sample of undergraduate students (Ruzibiza et al., 2018). On the other hand, invoking resilience as a mediator between childhood adversity and poor psychological outcomes implies the existence of a direct effect of early traumatic experiences on resilience itself, making low resilience a risk factor (Kraemer et al. 2001) for stress-related outcomes. More in general, invoking resilience as a mediator assumes that a given independent variable could exert an effect on resilience itself. This assumption suggests that resilience is a dynamic and changeable constructs (Stainton et al., 2018), hence several factors that could promote fluctuations of resilience over time should be taken into account in resilience research.

Our results are in line with the view that both personal and contextual resilience are affected by a risky family environment: in particular, personal assets may be weakly affected by environmental factors, while showing a large impact on PLEs, carrying the largest part of the mediated effect on PLEs.

In our model, the effect of risky family environment on Contextual Resilience could be due to partial overlapping of the contents of the two constructs. Social resources, on the other hand, have been confirmed to act as moderators by one of the few longitudinal studies available (Crush et al., 2018; Riches et al., 2019). The presence of good social resources, in fact, could be spared from the destructive effects of a stressful familial environment. Nevertheless, in order to access environmental social resources, one needs to be socially competent in the first place, a functional domain that has been confirmed to be impaired in individuals with PLE (Chisholm et al., 2018). Social competence is a particular factor in resilience as assessed by RSA. Inspecting our CFA, social competence has the lesser loading on personal resilience, while in our path model modification indices required adding a covariance (0.3) between social competence and social resources, indicating that social competence and social resources are the links between personal and interpersonal resilience.

The role of resilience as a protective factor could be questioned as the link between a harsh familial environment and low personal resilience suggests that early adversity may hinder the development of functional personal resilience resources. These in turn may play a role in the development of PLEs, for example failing to protect the individual from recent stressors (Bhavsar et al., 2019).

One model that could involve both a moderator and mediator role of resilience could be a dual-stage one, in which early traumatic experiences negatively affect resilience in the first place, and resilience in turn moderates the response to subsequent traumatic events.

This study opens important questions about the factor structure of resilience in different populations, namely help-seeking populations such as At Risk Mental States, Ultra-High risk samples and First Episode Psychosis. A number of factors could affect the latent structure of resilience in such populations, so exploring this issue in further studies could be of great relevance.

## Limitations

Our study has some conceptual and methodological limitations.

Firstly, Risky Family Questionnaire (RFQ) was not previously validated. In order to address this issue, we have performed both EFA and CFA. It worth noting that RFQ focuses on dysfunctional familial environment rather than childhood traumatic experiences in the broadest sense.

Secondly, some statistical limitations are present on the RSA CFA: the two-dimensional model has been particularly difficult to converge, and several interventions of the initial values were made in order to reach convergence. This is possibly due to the fact that only two factors were loaded onto “Interpersonal Resilience”. Despite having convergence issues, we chose not to re-specify the model because we aimed at specifying a soundly theory-driven model in the first place.

Finally, some of the interpretations we have provided in the discussion may be flawed by the cross-sectional design of this study. When addressing traumatic experiences using a retrospective self-report instrument, as the vast majority of studies actually do, recall bias is a major concern that could be eliminated only by a longitudinal non self-report design.

## Conclusions

The main findings of this study suggest that resilience could be considered two-dimensional construct. Personal resilience, as opposite to contextual resilience, shows a mediating effect within the putative causal pathways from risky family environment to PLE. These evidence could have a clinical relevance as they suggest that personal and contextual resilience factors should be addressed separately in the context of early of early interventions, and could represent an important therapeutic target in subjects exposed to childhood adversity. One possible extension of our findings, in future studies, could be in assessing the differential impact of early and later traumatic events in both general and clinical populations towards PLEs, as well as a dual mediation/moderation model. Under a clinical perspective, exploring in detail resiliency resources could help to a better design of individualized tailored treatment plans in young help-seeking individuals with a history of risky family environment.

## Data Availability

data available on request by the authors

## Acknowledgments

The authors would like to thank all the participants to the study. No funding was received for this study.

